# Adverse sequelae of the COVID-19 pandemic on mental health care in six low- and middle-income countries: MASC study

**DOI:** 10.1101/2024.06.18.24309132

**Authors:** Charlotte Hanlon, Heidi Lempp, Atalay Alem, Azeb Asaminew Alemu, Rubén Alvarado, Olatunde Ayinde, Adekunle Adesola, Elaine Brohan, Thandi Davies, Wubalem Fekadu, Oye Gureje, Lucy Jalagania, Nino Makhashvili, Awoke Mihretu, Eleni Misganaw, Maria Milenova, Tamar Mujirishvili, Olha Myshakivska, Irina Pinchuk, Camila Solis-Araya, Katherine Sorsdahl, Gonzalo Soto-Brandt, Ezra Susser, Olga Toro-Devia, Nicole Votruba, Anuprabha Wickramasinghe, Shehan Williams, Graham Thornicroft

## Abstract

**Objectives:** In this context the MASC study aimed to: (1) identify the consequences of the pandemic for mental health services and people with pre-existing mental health conditions (MHCs) in 6 low- and middle-income countries; and (2) identify good practice to mitigate these impacts.

**Design:** An observational study, using a mixed-methods convergent design triangulating data from (1) semi-structured interviews or focus groups and/or a self-completed survey; (2) routine service utilization data; (3) local grey literature; and (4) expert consultation.

**Setting:** The study was conducted in Chile, Ethiopia, Georgia, Nigeria, South Africa and Sri Lanka.

**Participants:** 121 key informants.

**Results:** We found clear evidence in all sites that the pandemic exacerbated pre-existing disadvantages experienced by people with MHCs and led to a deterioration in the availability and quality of care, especially for psychosocial care. Alongside increased vulnerability to COVID-19, people with MHCs faced additional barriers to accessing prevention and treatment interventions compared to the general population. To varying extents, sites showed accelerated implementation of digital technologies, but with evidence of worsening inequities in access. Where primary care-based mental health care was more developed or prioritised, systems seemed more resilient and adaptive.

**Conclusion:** Our findings have the following implications. First, mental health service reductions are clear examples of ‘structural stigma’, namely policy level decisions in healthcare which place a low priority upon services for people with MHCs. Second, integration of mental health care into all general health care settings is key to ensuring accessibility and parity of physical and mental health care. Third, digital innovations should be designed to strengthen and not fragment systems. We discuss these findings in terms of anticipating such challenges in future and preparing layers of resilience.

**Methodological Strengths and Limitations of this Study:**

- The MASC study used a **mixed-methods convergent design**, incorporating both quantitative and qualitative data, which allowed for a nuanced understanding of the COVID-19 pandemic impact on mental health services across multiple countries. This design enabled our study to triangulate data capturing both numerical trends and in-depth perspectives, increasing the validity and reliability of the findings.
- The study covered **six diverse low- and middle-income countries across five continents**, enabling cross-country comparisons and insights into varied socio-cultural and economic contexts. This geographic diversity strengthens the study’s generalisability to other settings with similar resource constraints.
- A mixture of **online and in-person data collection in local languages** was employed, overcoming pandemic-related barriers and enhancing accessibility for participants across the regions, which minimised data collection disruptions.
- The study faced **limitations in obtaining comparable quantitative data across countries**, with concerns about the accuracy and reliability of routine data.
- Reliance on **national experts may have introduced potential biases** if experts were not fully representative of the wider mental health context.

## Introduction

Prior to the global COVID-19 pandemic, mental health care was delivered to a small minority of people in the world with mental health conditions (MHCs)^1^. For people with severe depression, for example, only 22%, 11% and 4% of those in high-, middle- and low-income countries respectively, received minimally effective treatment^2^. During the early phases of the COVID-19 pandemic data suggested that in many countries the incidence of some types of MHCs, especially anxiety and depression, increased at the population level^3 4^. At the same time, mortality rates from COVID-19 infection were found to be higher among people with MHCs than among the general population^5^. During this initial period, evidence emerged that the COVID-19 crisis response in many countries had the effect of weakening mental health services and systems^6^. It became clear that the mental health targets set by the United Nations Sustainable Development Goals aim to ‘Leave No-one Behind’ were at risk from a degradation of mental health care^7^.

The *Mental health care: Adverse Sequelae of COVID-19* (MASC) project was framed by the concept of the ‘mental health treatment gap’. This refers to the proportion of people in any community who need evidence-based mental health treatment who receive it^8^. In low- and middle-income countries (LMICs) over 80% of people with severe MHCs receive no mental health services^9^. This contributes to persistence of symptoms, health deterioration, ostracism^10^, long-term disability, exclusion from the workforce, social isolation, poorer physical health and premature mortality^11 12^. In LMICs, mental health conditions account for 7.4% of the global burden of disease^13^, but only 0.5% of these countries’ health budgets are spent on mental health care. Community mental health care is scarce, specialists are in short supply, and services are mostly hospital-based^14^.

Numerous studies have examined COVID-19 in relation to the psychological wellbeing of the general population, or of the health workforce, with relatively less focus on people with pre-existing MHCs and mental health services^15^. Evidence from mostly high-income countries indicates that a combination of factors related to the pandemic itself, and to the prevention and mitigation strategies, were responsible for infringement of the right to mental health of people with MHCs, with increased inequities in comparison with the general population^16^. Evidence of negative impacts of lockdown on people with existing MHCs and the adequacy of mental health services were reported from Switzerland^17^, Italy^18^ and Norway^17^.

Reductions in the availability and quality of mental health services were reported from Spain^19^ and significant unmet needs of service users, including inability to access welfare benefits were reported in the USA^20^. General adverse impacts of the pandemic were also reported: in Sweden 20% of people with pre-existing MHCs reported an increase in their psychiatric medication compared to pre-pandemic^21^. In a multi-centre study from Austria, Denmark and Germany, people with bipolar disorder reported an increase in negative lifestyles, including greater use of alcohol and smoking, and an increase in boredom, depression, somatization, anxiety, distress due to social distancing, and poorer sleep quality^22^. Impacts of COVID-19 on the physical health of people with severe MHCs were clear; including higher rates of COVID-19 infection, hospitalization and mortality for people with a pre-existing severe MHC^23 24^. Nonetheless, lack of prioritisation or explicit exclusion of people with severe MHCs from COVID-19 vaccination programmes was observed in many settings^25 26^.

There have been notably fewer studies from settings in LMICs to examine impacts of COVID-19 on mental health services and people with MHCs; most have been conducted in a single setting in a single country. In Indonesia, the number of people with MHCs who were shackled increased from 5,200 in 2019 to 6,200 in 2020^27^. In China, the prevalence of post-traumatic stress disorder (PTSD) was high among people with MHCs^28^. In India, most people with MHCs (72.6%) reported a positive impact of the pandemic due to the increased availability of family support^29^. However, some (22.6%) stopped medications, many had difficulties accessing health services and experienced increased interpersonal conflict, sleep difficulties and a surge in screen time.

While the COVID-19 pandemic undoubtedly drove innovation in responding to population mental health needs globally^30^, there have been no cross-country studies examining the pattern of responses and impacts of mental health service changes on people with severe MHCs (such as psychotic disorders and bipolar disorder) in LMICs.

In this context, the aims of the MASC study were to: (1) identify the consequences of the COVID-19 pandemic for mental health services and people with pre-existing MHCs in six LMICs; and (2) identify examples of good practice in efforts to mitigate these impacts in the future. Country level findings from this dataset have been presented for Chile^31^, Ethiopia^32^ and South Africa^33^. In this paper we report a comparative analysis of the main cross-country findings of the MASC study, focusing on cross-country similarities and differences in impacts and responses to inform future preparedness and system resilience.

## Materials and methods

The MASC project was conducted in six LMICs in five continents (Chile, Ethiopia, Georgia, Nigeria, South Africa and Sri Lanka). This was an observational study with a mixed-methods convergent design. Details of the study sites and the data collection and methods employed are shown in Table 1.

**Table 1.** Data gathering methods and samples across the 6 countries of the MASC study.

|  | **Chile** | **Ethiopia** | **Georgia** | **Nigeria** | **South Africa** | **Sri Lanka** |
| --- | --- | --- | --- | --- | --- | --- |
| Time of data collection | 01/04/2021-30/04/2021 | 01/02/2021-28/02/2021 | 15/12/2020-31/01/2021 | 01/06/2021-30/06/2021 | 01/07/2021-30/09/2021 | 01/05/2021-30/06/2021 |
| Researchers conducting the qualitative study | 2 (OT, CS)<br>Both female;<br>1 mental health professional, 1 mental health researcher<br>Qualifications: PhD, MSc | 2 (AM, WF)<br>Both male;<br>1 clinical psychologist, 1 mental health professional;<br>Qualifications: MSc, MA | 2 (TM, LD)<br>Both female;<br>Both Mental Health professionals;<br>Qualifications: PhD Student; MA student | 2 (OA, AdA)<br>Both male;<br>Both mental health professionals;<br>Qualifications: psychiatrists | 2 (TD, KS)<br>Both female:<br>Registered counsellor, health psychologist<br>Qualification: both PhD | 2 (AW, SW)<br>1 female, 1 male;<br>Both psychiatrists<br>Qualifications: MD, MSc |
| Location of interviews/focus groups | Online | Clinical settings, home or office and online | Online and face - to-face interviews | Online meeting and telephone interviews | Online | Online meeting or clinical setting |
| Language (and translated into English) | Spanish | Amharic | Georgian | English | English | English or Sinhala |
| Recruitment sample type | Purposive | Purposive and snowballing | Purposive | Purposive | Purposive | Purposive |
| Average duration of focus groups or interviews | 1-1.5 hours | 40 minutes | 1-1.5 hours | 45 min – 1.5 hours | 40 minutes | 45-60 minutes |
| Number of participants and roles | 28 participants (18 women, 10 men) in 3 focus group discussions: | 11 expert group<br>18 participants in qualitative interviews (3 | 10 Expert Group members* (9 women, 1 man), each also participated in qualitative | 15 participants (6 women, 9 men) completed the key informant interview. | 10 expert group members<br>17 stakeholders (10 women, 7 | 26 stakeholders participating in interviews and completing quantitative tool (18 men, 8 |
|  | 11 health services professionals: 5 working at the primary health care level, 3 from community mental health centres, 3 from psychiatric services in general hospitals. 10 Policy makers: 2 from the ministry of health, 8 mental health care network directors. 7 experts by experience: independent users and family organizations | women, 15 men): individuals with mental health conditions (n=4), representatives from national bodies involved in the mental health response to COVID-19 (n=2), mental health experts (n=8), a religious leader (n=1), a human rights advocate (n=1), and civic society representatives (n=2). | interviews and 7 completed narrative tool: Inpatient service providers - 2 persons Outpatient ambulatory service providers - 3 persons Community-based Service provider (Assertive Community Team) - 1 person Community-based MH Service (Mobile Team) - 3 persons Community-based Service (Crisis Intervention Centre) - 1 person Non-governmental (NGO) mental health service - 3 persons Psychosocial Centre - 1 person Service user (person with lived experience) from a local NGO - 1 person | 12 participants (5 females and 7 males) took part in the online expert panel meetings.<br>Psychiatrists working in different care settings, psychiatrists with administrator roles, medical officers at different levels of care, nurses, programme officers and managers in NGO settings, Social Workers, Psychologists, Service users and family care givers | men) (12 interviews; 9 narrative tool and quantitative survey): public sector servants, NGO service providers, psychiatrists, psychologists, mental health nurses, counsellors and occupational therapists. (no service users/carers) | women): Health Service Professionals=14 (Medical officer mental health, community psychiatric nurses, Social Worker, health education nurse, Consultant Psychiatrists of different settings like forensic and general adult, pharmacist)<br>Policy makers, stakeholders and NGOs=7 (chairperson of Befrienders, members of civil society, director and deputy director of National Institute of mental health, Sri Lanka, other administrators, Chairman of the Board of studies - Psychiatry, Sri Lanka) |
|  |  |  | Human rights advocate, NGO representative - 1 person<br>Addiction service provider - 1 person |  |  | Expert by Experience and Advocates=5 (elders' home manager, substance counsellor, care giver, mental health advocacy group, consumer action forum founder) |
| Software used | No | Opencode | No | MAXQDA | Opencode | Opencode |
| Number of researchers involved in coding | 2 | 2 | 2 | 2 | 2 | 4 |
| Regions | North, Centre, South Centre, South and Austral Macrozones | rural and urban settings in Ethiopia | Tbilisi, Batumi, Rustavi | 6 geopolitical zones of Nigeria | Western Cape Province | Sri Lanka (Western province, North central province, other) |

\*Number for roles held by stakeholders is more than 10 because some stakeholders hold multiple roles.

The quantitative component compared service utilisation data from public mental health facilities between 2019 and 2021 from the available health management and information system statistical registries of the local, regional or national health services in the available study sites and countries. Analyses of country level service utilisation data have been reported previously^31–33^ but our analysis focused on patterns across settings and incorporated additional data from Georgia, Nigeria and Sri Lanka. Given the heterogeneity across sites in terms of the availability of in-patient/out-patient data and the service level within the health system, the findings were analysed descriptively, presented in graphical form and combined with key informant reports of the COVID-19 pandemic impacts on mental health services at all levels of the health system.

For the qualitative component, we conducted semi-structured focus groups and/or interviews with purposively selected key informants, including mental health services providers, planners, decision-makers, and service users and members of relevant organisations in each country. Potential respondents were approached by phone or email and interviews were conducted virtually or in-person depending on the setting. Written, informed consent was obtained from participants in all countries. 121 people participated in the qualitative study, with 16 refusals in total across the sites. In Chile, a master’s student was present in addition to the interviewer but in other sites just the interviewer was present. See Supplementary File 1 for the topic guide, which explored impacts of the pandemic on people with MHCs in the community; impacts on mental health care availability and quality at the primary, secondary and tertiary level; access to physical health care; policies, plans and organisational level impacts on people with MHCs and services. See Supplementary File 5 for the COREQ checklist for reporting qualitative studies. In each country, researchers who carried out data collection had experience and training in qualitative research. Researchers had pre-existing professional links to some respondents but not clinical relationships. They were known as researchers on mental health or mental health clinicians in their countries.

Interviews were conducted in local languages, transcribed and translated into English. Country level analyses were first conducted by country teams using template analysis^34^. This approach centres on the set up of a finalised coding template that includes the themes identified by the researchers as relevant in the qualitative data set and arrange these in a meaningful way^35^. The analytical process begins with the definition of *a priori* themes, subthemes and codes; and as the analysis proceeds, these may be revised or disregarded if they do not relate to the empirical data. A cross-country analysis took place, with each site contributing key findings for the main themes/sub-themes and illustrative accounts.

We also used a quantitative rating tool (see Supplementary File 2) to seek structured input from additional key stakeholders. The tool included reasons for change in service utilisation and mitigation strategies. Alongside this blueprint, country teams identified relevant published or grey literature from each country, including policies, plans and programmatic reports. Where possible, a specially convened national expert group oversaw the study, guided identification of key informants and relevant grey literature, and reviewed the emerging findings. The results were integrated through triangulation^36^ using a convergent coding matrix, to identify key results for each predefined theme. See Supplementary file 3 for the cross-country matrix. Data underlying quantitative health service utilisation data are included in Supplementary file 4. For ease of references the countries are coded as follows: CH: Chile; ET: Ethiopia; GE: Georgia; NI: Nigeria; SA: South Africa; SL: Sri Lanka).

### Patient and public involvement

People with lived experience of mental health conditions were included in the national expert groups, involved in review of triangulated data sources. In Ethiopia, a mental health service user representative (co-author EM) was involved in analysis, write-up and dissemination.

### Ethical considerations

Ethical approval was obtained from King’s College London Research Ethics Committee (HR-20/21-21056); Chile: Institutional Review Board of the Faculty of Medicine of the University of Chile (project No. 270-2020); Ethiopia: Institutional Review Board of the College of Health Sciences, Addis Ababa University (Ref 087/20/CDT); Georgia (Ethics Committee at Ilia State University; Ref. IL20/12/11.3.); Nigeria: University of Ibadan/University College Hospital Ethics Committee (UI/EC/20/0366); South Africa: University of Cape Town Human Ethics Research Council (Ref: 552/2020); Sri Lanka: Rajarata University of Sri Lanka Ethics Review Committee (ERC/2021/06).

## Results

Pervasive negative impacts of the COVID-19 pandemic on people with pre-existing MHCs were reported across the six countries, affecting life in the community and closely linked to difficulties with access to adequate health, psychological and social care. Mental health-related stigma amplified adverse consequences of the pandemic.

### Life in the community

Respondents across all countries reported how pandemic control measures and economic disruption had adversely affected people with MHCs and their families, exacerbating pre-existing disadvantage.

### Stigma and human rights violations

Stigma towards people with MHCs increased in some (CH, ET, NI, SL) but not all countries. Pre-existing perceptions of the unreliability and dangerousness of people with MHCs combined with fear of contagion of COVID-19 to magnify social exclusion (CH, ET, SL).

Conflating COVID-19 infection with mental illness fuelled stigma [NI]:

> *‘… when the people see that the person has a relapse, they start to think the COVID has entered the person’s brain… they call it madness, not mental disorder, which further worsen the stigma. The person has COVID which is an infectious disease… it’s like having double burden which is a worse situation for somebody with a mental health condition’*.

> *Psychiatrist, Nigeria*

Public and family-level stigma was exacerbated by the deterioration in mental health of some people with MHCs (ET, NI, SA). Worsened mental health led to families resorting to use of physical restraints for unmanageable behavioural disturbance (ET, NI). Although increased public discussion about mental health in some countries (ET, NI, SA) raised awareness and reduced stigma, this was largely in relation to depression and anxiety.

### Economic and social life

Aggravation of economic problems was reported to have differentially negatively affected people with MHCs (CH, ET, NI, SA), for example, being laid off first or at higher risk of unemployment because they were seen as less reliable workers (ET, NI).

> *‘Some service users were laid [off] from their work due to COVID-19 and some of them still didn’t get a job because once they became out of the system, it has been difficult to be re-employed since the macroeconomy is weak to accommodate many people.’*

> *Mental health service user, Ethiopia*

Access to public funds for economic support and food supply for people with MHCs were either difficult to access (CH), limited (NI, SA, SL) or non-existent (ET, GE). Reduced social support and increased isolation of people with MHCs were reported across all study countries. The reasons included suspension or disruption of in-patient, social care and community services (CH, ET, GE, NI, SA), extensive quarantines, enforced home-based isolation, and social distancing measures in all countries.

> *‘People with mental illness already had difficulty communicating and integrating with society, and in this setting [the pandemic] their situation became even worse.’*

> *NGO Representative, Georgia*

### Social care and residence

Increased vulnerability of people with severe MHCs to homelessness was reported in several countries (CH, ET, NI, SA) due to the worsening economic climate (NI), restricted movements (SA), community residential homes being unable to maintain services for the poorer community members (ET, SA), religious and traditional healing sites being unable to provide shelter (ET, NI, SL), families being overwhelmed because of the lack of access to care and other support (ET), and overcrowding at home (CH). However, in Chile, newly established hostels for homeless people which included support from mental health teams were reported. In Sri Lanka, care homes may have helped to mitigate the risk of homelessness. People with MHCs living in social/care homes were particularly vulnerable to COVID-19 outbreaks and experienced social isolation due to family visiting bans (SA, SL).

### Health systems, governance, and legislation

#### Legislation or policies affecting people with mental health conditions

While there were no reports of discriminatory policies or laws introduced because of the pandemic, there was evidence of a lack of policies designed to uphold the rights of people with MHCs within COVID-19 responses (ET, GE, SA). In some places, this absence rendered people with MHCs vulnerable to coercive practices (ET, NI) or exclusion from care (SA):

> *‘Nobody had given any thought to what would happen in the psychiatric hospitals. We were just tasked to find a way… Unlike everywhere else in the health system where special provision had been made for people coming in and needing medical attention, nothing was done for people with mental illnesses.’*

> *Psychiatrist, South Africa*

Sri Lanka had strong policy commitment to maintain mental health care, evident in written documentation (three special circulars issued through the health ministry) and in its implementation. In other countries (ET, NI), national level commitments were not replicated at sub-national levels or ignored the needs of local services (CH) or were not associated with concrete action to implement mental health recommendations as part of the pandemic response (ET, GE, NI, SA).

### Co-ordination of COVID-19 response in mental health services

Overall, there was a lack of preparedness for an adequate pandemic response in mental health services in all sites, particularly for remote delivery of care. In some countries, COVID-19 protocols for mental health services were delayed (ET), variably implemented (ET, NI), left to facilities and service providers to develop (GE, SA) or could not be implemented without additional resources (ET). Responses were notably better in Sri Lanka

### Resourcing and programming of mental health care

Although mental health services were classified as essential services in most countries (CH, ET, SA, SL), in practice this did not materialise except for Sri Lanka. COVID-19 exposed and exacerbated the pre-existing poor resourcing of the mental health sector.

> *‘I think that historically mental health has always been under-catered for. I think that right now, all the disciplines are taking a cut. And that cut is happening at best proportionately. But we have always been underserviced and now they are taking the same amount away from everyone which means that we are going to feel it even more than everyone else. ‘*

> *Psychiatrist, South Africa*

When the pandemic arrived, some government mental health care budgets were diverted to pandemic response (ET, SA) or maintained but found to be inadequate in the face of increasing demand (CH), but other countries protected (SL) or even increased (GE: 5% increase in 2021) their budgets or benefited from external funding (NI, SL). In SL, World Bank funding supported services for mental health rehabilitation, people with developmental disabilities and development of COVID-19 wards at the National Institute of Mental Health, for mental health nurse training, and a mental health helpline.

> *‘The allocation of mental health resources to COVID-19 implies that people with mental health conditions were not getting the required services. There are other hospitals and treatment centres for different specializations such as orthopaedic, internal medicine or general health facilities, but their resources were not taken. … Why mental health care?’*

> *Psychiatrist, Ethiopia*

Plans to expand access to mental health care through training of primary health care workers were put on hold in some countries (ET) but accelerated in others (GE). Structures for supervising task-shared mental health care were disrupted (ET, NI, SA). Across countries, a paucity of routine data on mental health system functioning and population mental health need was reported and undermined both preparedness and response.

### Impacts and adaptations from mental health services

#### Access and availability of mental health services

In four of the six countries (ET, GE, NI, SA), there was limited pre-existing integration of mental health care within primary care, which constrained options for making mental health care locally available when the pandemic began. When present, mental health care in primary care was not always prioritised as an essential service (ET, SA). Sri Lanka was an exception, due to extensive pre-existing integration of mental health in PHC that was a legacy of a previous humanitarian crisis - the 2004 tsunami. Even so, periodic disruptions occurred due to infection waves and lockdowns. In Georgia, rapid expansion of capacity strengthening for delivery of mental health care in primary care was rolled out as a priority response. In South Africa, community health workers linked to primary care made efforts to deliver medication to the homes of people known to have a severe MHC.

Mental health care in general hospital settings was either very limited pre-pandemic (GE, ET) or became less available (CH, NI, SA), further increasing reliance on centralised specialist services.

Mental health services and availability of specialists at all levels in the health system were disrupted by movement restrictions affecting staff (NI), lack of personal protective equipment (NI), redeployment of staff to COVID-19 clinical duties (CH, ET, SA, SL) and COVID-19 related illness or quarantine of staff (CH, SL).

> *‘There was no reduction or redeployment of healthcare staff, but there was an increase in staff taking paid sick leave, and this has impacted the patients who were no longer receiving adequate services and care [due to staff shortages], for example, a psychologist who would have otherwise continued with psychotherapy, or any needed interventions.’*

> *Health Service Professional, Chile*

Tertiary mental health services were not spared disruption. Reassignment of specialist mental health facilities and in-patient wards to COVID-19 activities occurred in some (ET, GE, NI, SA) countries. In Sri Lanka this was small-scale and only during the peak of the pandemic. Attendance for out-patient mental health care was discouraged (SA), triage systems were introduced to prioritise emergency presentations (NI, SA) and the intervals between appointments was lengthened (ET, NI, SA).

> *‘The Mobile Team service was suspended for some time because home visits were dangerous for both patient and their family and mobile teams as well.’*

> *Community mobile team service provider, Georgia*

The number of in-patient beds for mental health care was reduced in some countries (ET, SA, SL), new admissions were suspended in others (CH, SA) or higher thresholds for admission were applied (CH, NI, SL, SA). In addition, admissions were suspended or reduced whenever there were outbreaks of COVID-19 on the wards (ET, GE, SL).

#### Utilisation of mental health services

There was markedly less use of public sector outpatient mental health services in five of the six countries (CH, ET, NI, SL, SA), although increased attendance occurred at private sector care in some settings (NI) and increased demand for emergency admission in others (CH, SA).

Compared with 2019 (pre-pandemic), in 2020 (first year of pandemic) there were clear reductions during all or part of 2020 for inpatient service use in ET, SA and SL (Fig 1).

**Fig 1:**
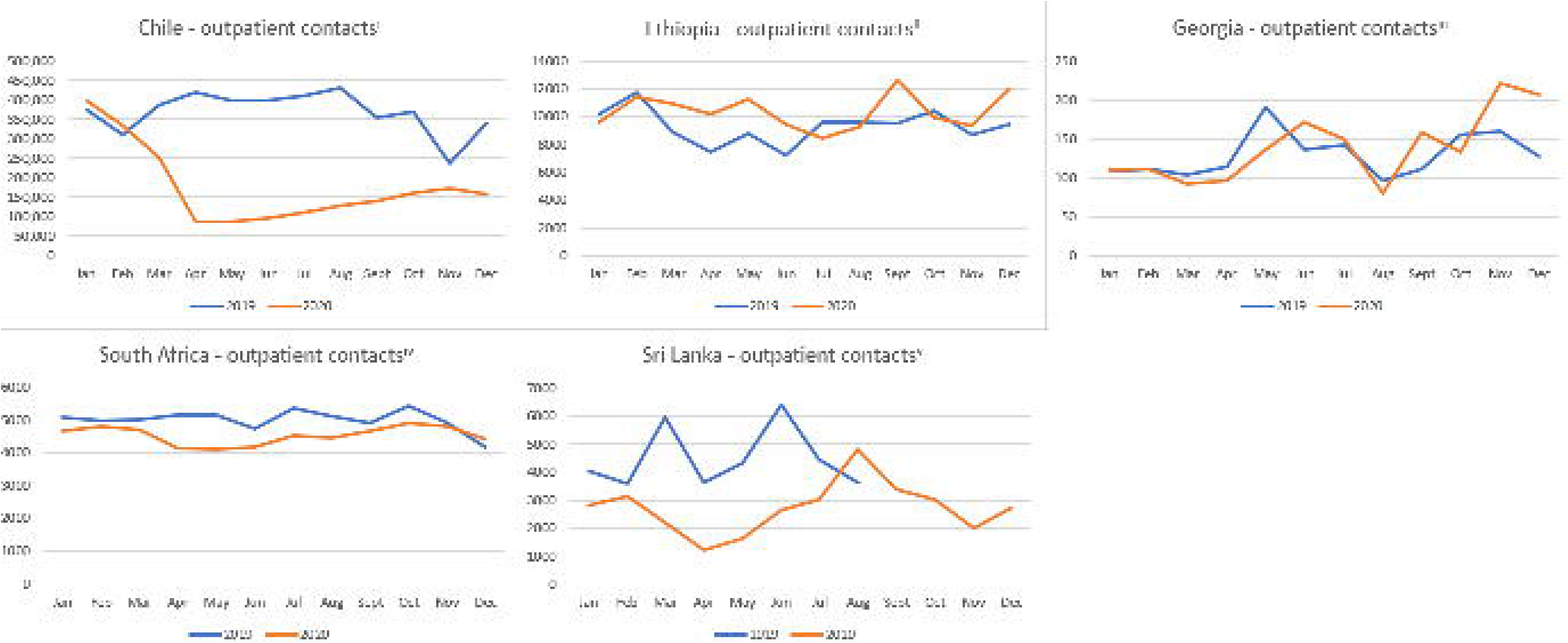
Mental health in-patient service utilisation in MASC countries in 2019 and 2020. ^a^Data from the national referral hospital in Addis Ababa, Ethiopia; ^b^National data, Sri Lanka; ^c^Data for the Western Cape, South Africa;

Regarding out-patient service utilisation there was a more mixed picture (Fig 2). There were persistent reductions throughout 2020 in out-patient use in some (CH, SA, SL) but not all (ET, GE) countries. In Ethiopia, maintained levels of out-patient contacts at the national referral hospital reflected the closure of other specialist out-patient clinics and, therefore, masked a de facto fall in per capita utilisation.

**Fig 2:**
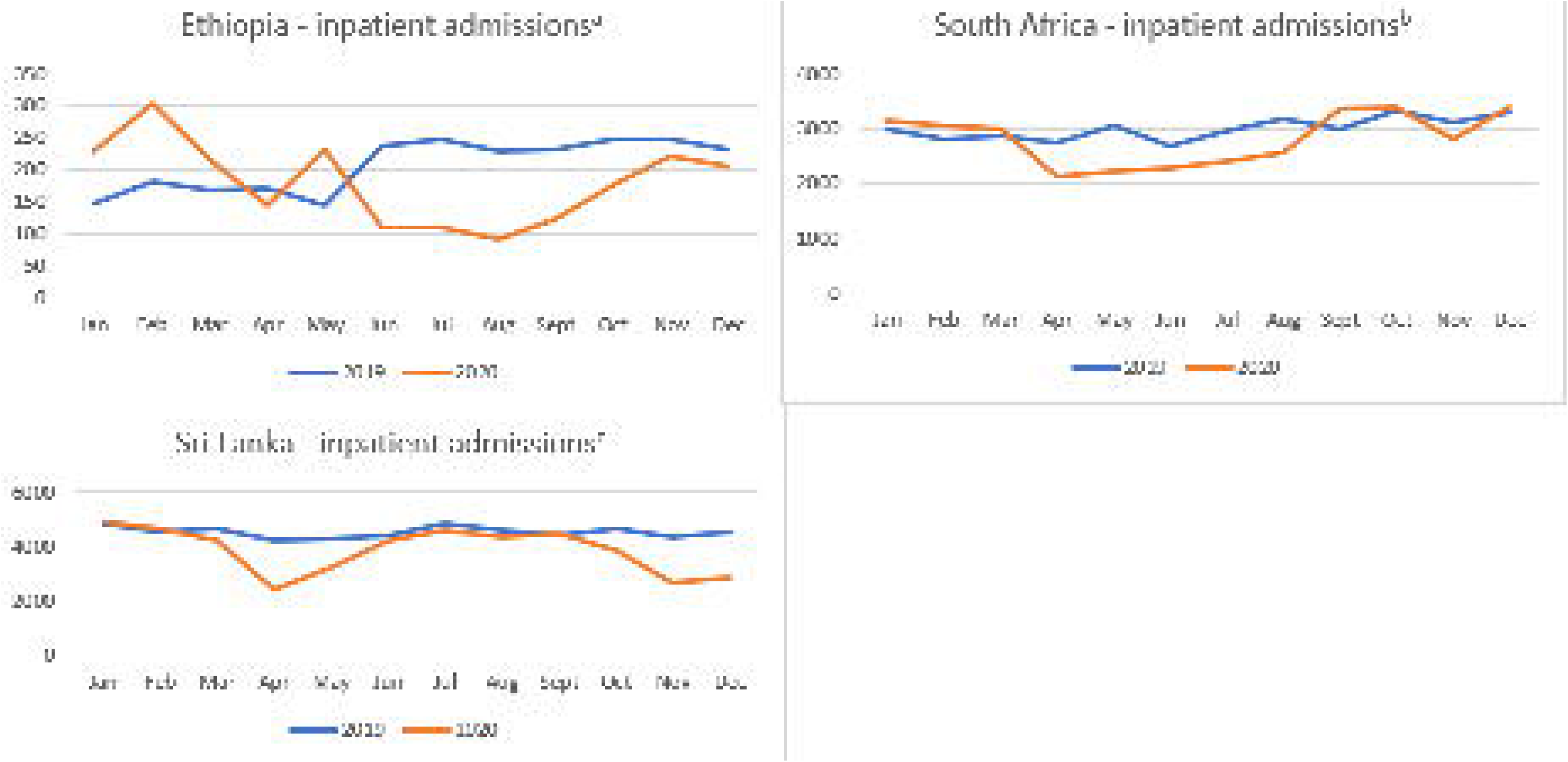
Mental health out-patient service utilisation in MASC countries in 2019 and 2020. ^i^*National data, Chile;* ^ii^*Data from the national referral hospital in Addis Ababa, Ethiopia;* ^iii^*Data from cities of Tbilisi, Batumi, Rustavi, Georgia;* ^iv^*Data for the Western Cape, South Africa;* ^v^*Data from Anuradhapura Teaching Hospital, Sri Lanka;*

Difficulty with transport to access centralised, specialist mental health care was problematic in all countries, exacerbated by movement restrictions. Attendance for mental health care was also reduced due to fear of infection (ET, NI, SA, GE, SL).

> *‘…Access to care generally was reduced drastically for some of the reasons I have mentioned earlier that the lockdown affected free movement of people across … and mental health services are not readily available within primary care, so many individuals needed to travel for sometimes 50 kilometres, 100 kilometres to be able to access mental healthcare services…’*

> *Psychiatrist, Nigeria*

#### Quality and adequacy of mental health care

Mental health care at all levels in the health system became more narrowly biomedical in all countries and was difficult to maintain, with increased costs and interruptions to supply of essential psychotropic medication in some countries (ET, NI, SA) and periodic stockouts in others (CH, GE). Most countries sought to find ways to allow continuity of medication supply for people with MHCs, including through home delivery (CH, SA, SL), sending prescriptions to more locally accessible pharmacies (ET) and writing prescriptions of longer duration (ET, SA).

Participation of service users in ensuring quality of primary and community mental health care ceased in Chile. Community-based psychosocial interventions, counselling, group workshops and community activities barely existed pre-pandemic in Ethiopia and Nigeria. In other countries, these were substantially reduced (CH, SL, GE) or stopped altogether (SA). In Georgia, pressure for mental health services in primary care increased, but health workers were ill-equipped to deliver mental health care, leading to concerns about quality of care. Supervision of primary care staff delivering mental health care was reduced in Chile, Nigeria and Sri Lanka.

> *‘Mental health services are maintained; however, psychosocial interventions and community activities have been suspended. The pharmacological treatment was followed up, and any additional psychosocial support service users needed were abandoned.’*

> *Health Service Professional, Chile*

In secondary and tertiary level mental health care, most countries saw increases in waiting times and significant reductions in consultation duration and frequency (all sites). Although new patients were assessed face-to-face in most cases, follow up appointments were replaced by issuing repeat prescriptions unless there was a clear clinical need for in-person review (ET, SL). The use of facemasks limited patient-doctor non-verbal communication and rapport (CH, ET, NI, SL, SA) and disease control measures reduced involvement of families in consultations (SL). This was later mitigated through use of transparent screens (SL, tertiary setting in ET).

In countries where psychological therapies in secondary and tertiary care were more widespread, there was a substantial reduction in availability (CH, ET, NI, SA, SL). This was due to lower prioritisation of this aspect of care (not considered essential) and the restrictions on face-to-face interactions (ET, NI, SL, SA). A further contributing factor was the redeployment of psychologists to support frontline health workers (CH, ET, GE). Availability of psychological interventions in Ethiopia, South Africa and Sri Lanka did not return to pre-pandemic levels even when disease control restrictions were eased. Group therapies, for example for people with substance use disorders, stopped entirely in some countries (ET, SA, SL). In three countries CH, GE, SA), efforts were made to transition psychological interventions to online platforms, but the inadequacy of this approach compared to face-to-face meetings was reported.

> *The Psychosocial Rehabilitation Centre for persons with severe mental illness closed at the end of March 2020, opened temporarily in September, but closed again in November due to the threat of the virus. This service was important for the patients, they visited it [the venue] for socialization and to communicate with each other, therapies were conducted, the environment was warm-hearted and comfortable*.

> *Psychosocial service provider, Georgia*

In-patient services were often suspended, or minimised, and patients were discharged earlier than usual (CH, ET, NI, SA, SL), with concern that the discharge was premature (ET, SA), but in other settings in-patient stays were prolonged due to staff shortages (GE). In-patients were negatively affected due to the restriction of family visits (CH, SA, SL).

### Transition to remote care

Many countries introduced remote care by phone or online in public sector services (CH, GE, SA, SL), but this was largely restricted to the private sector or non-governmental organisations in ET and NI. Phone-based activities included clinical assessments and follow-up appointments (CH, NI, SA), responding to queries from people with MHCs and their caregivers (GE, SL), and delivery of psychological therapies (CH, GE, SA).

The digital divide prevented widespread use of digital platforms for psychological interventions in CH, ET, SA and NI.

> *‘…. we have come across [low awareness of remote psychological services] in some important way, not only with the digital illiteracy of our [service] users but also with our own digital illiteracy, using our own equipment and that this [situation] has meant a significant gap and access to technologies … It [the situation] has to do with the geography, with the countryside [and] with the connectivity of some areas…’*

> *Health Service Professional, Chile*

No resources were allocated to the transition to phone/internet-based care, with costs falling to providers and adverse consequences for quality (CH). Poor training and familiarity with this mode of consultation by health workers was an additional barrier (NI). Barriers to access were seen for elderly, rural communities, and very poor families, fuelling concerns that those most in need were least likely to be able to access digital mental health care (CH, GE, SA). This led to early resumption of face-to-face consultations for high-risk cases (CH). Privacy and confidentiality in remote care were concerns (CH, SA), especially for children and adolescents.

> *‘It’s just that people don’t always have phones, you can’t always get hold of them. And also, people don’t always have access to private spaces to speak to the mental health nurses in their homes. So, it just became difficult; the technology did not help hugely.’*

> *Psychiatrist, South Africa*

#### Physical and mental health

COVID-19 exposed systemic difficulties in the provision of health care for people with MHCs across six countries, apart from SA, in the various healthcare sectors (primary, secondary, social care or NGOs). During the pandemic there was limited, inconsistent or worsened access and delivery of physical health care, with inadequate preparation for the COVID-19 pandemic in already under-resourced services. People with mental health conditions or developmental disabilities overall seemed to experience direct or indirect discrimination from healthcare workers, e.g. stigmatising attitudes, fear of unpredictable or aggressive behaviour from health care workers in most countries (CH, ET, GE, NI, SL).

#### Information on COVID-19 and access to protective interventions

Although all countries provided information to the public about COVID-19, the information was either not easily accessible for many people with MHCs (ET) or was not tailored to their specific needs and concerns (CH, ET, GE, SA, SL). In some instances, information appeared to exacerbate mental ill-health and reduce uptake of protective measures because of frightening messaging (ET) or because of inaccurate and inconsistent information accessed via social media (CH, GE). In response to the need for clear information for people with MHCs and their caregivers, SL launched a country wide telephone line to address queries.

Telephone services were provided from the outpatient clinic and mobile team members in GE who provided educational instructions to the patients.

In most countries, people with MHCs from more vulnerable populations (homeless, international migrants) had limited or no access to personal protection from COVID-19. Some countries reported that people with MHCs were less able to afford protective interventions (e.g., masks, sanitiser or gloves) due to economic disadvantages (ET, GE, NI).

> ‘*Patients did not have soap and no disinfectant solution was available as it contains alcohol and the administration did not allow these liquids in the wards for fear that patients would drink it. No alternatives were used, such as alcohol wipes or soap. Patients did not wear a mask, even if the rules prohibited staying indoors without a mask’*.

> *Service User, Georgia*

In Nigeria, COVID-19 testing was by demand and, therefore less testing was conducted amongst those with severe MHCs.

There was some evidence of a mismatch between national policies prioritising people with MHCs for vaccination (ET, GE), and its implementation in communities. In some countries, this was partly due to low levels of awareness amongst people with MHCs (CH).

#### COVID-19 care for people with mental health conditions

Several countries reported that people with MHCs faced discrimination in accessing COVID-19 care and/or that the care they received was inferior to those without MHCs. Examples were provided of individuals with symptomatic MHCs being excluded from ambulance services or COVID-19 facilities (SL, ). People with symptomatic MHCs who had COVID-19 were admitted to mental health institutions for COVID-19 care in GE, SA and SL regardless of whether the person’s mental state warranted in-patient psychiatric care. In Chile, people with MHCs were excluded from hotels for quarantining people with COVID-19. People with MHCs in COVID-19 facilities reportedly received less attention, lower standard of care and were stigmatised by healthcare workers (ET) due to fear and lack of knowledge about mental illnesses.

> *‘If a psychiatric patient reacts negatively due to lack of oxygen in the emergency setting, health care providers thought the patient is psychotic. They associated all maladaptive behaviours of patients in the COVID-19 treatment with mental illness. These could be due to a lack of mental health knowledge and thus, stigmatizing attitudes. Thus, we have been doing some activities to increase the mental health knowledge of the healthcare workers.’*

> *Director of a hospital, Ethiopia*

Intensive care unit beds were reportedly accessible to people with MHCs if needed in Chile and South Africa, but with concerns about unequal access in Georgia and Sri Lanka. In Ethiopia access to intensive care facilities was extremely constrained for the whole population, with no reports of differential access. People with MHCs in Chile who lived in supported housing had less access to COVID-19 vaccines, despite officially being prioritised.

#### Access to physical healthcare

Longstanding discrimination pre-pandemic impeded receipt of care for non-COVID-19 physical health conditions in people with MHCs in Ethiopia and Nigeria, and to a lesser extent Sri Lanka. In all countries, the pandemic further disrupted access to physical healthcare, with no specific support extended to people with MHCs, and barriers encountered in some countries; for example, unaffordability of transport and care (ET, NI), exclusion from in-patient physical health care if mental health symptoms were apparent (GE), and fewer and shorter contacts with mental health professionals (ET, NI, SA) reducing opportunities to detect co-morbid illness.

> *We had a team of both physical and mental health care workers in the psychiatric ward before COVID-19. But during the pandemic, all medical professionals went for COVID-19 response. They were getting additional incentives, risk payment when they work at the COVID-19 response. No one was delivering the care for the comorbid physical conditions of our psychiatric patients. Only one medical professional remained to deliver the physical health care*.

> *Psychiatrist, Ethiopia*

People who were homeless, or who had alcohol or substance use disorders were excluded from physical health care in Chile.

#### COVID-19 protection and care within mental health and social services

Protections for people with MHCs and staff on in-patient wards were reportedly inadequate, due to lack of access to disinfectant (GE), protective personal equipment (ET, NI), or with respect to mask wearing (GE, SA). In some countries (ET, GE, SA, SL), in-patient facilities reported challenges with safely separating patients who were COVID-19 positive from other in-patients. Human rights violations were reported, with patients who tested positive apparently isolated with minimal social contact in ET and admission of COVID-positive patients to psychiatric wards (SA). It was difficult for families to obtain information about in-patients (CH). COVID-19 care for in-patients who tested positive was said to be poor in some countries (CH, ET), others opened dedicated wards (SL) and introduced rigorous quarantine procedures for newly admitted patients (SL). In residential social care settings in Ethiopia and Georgia, lack of COVID-19 testing or procedures to isolate those with suspected COVID-19 were apparent.

#### Innovations and strengthening system resilience

Examples of innovation and measures that strengthened health system resilience in response to the pandemic were evident in all countries. Key measures are summarised in Fig 3 in relation to: strengthening community supports; maintaining access to mental health care; quality of mental health care; and co-ordination of care.

**Fig 3.**
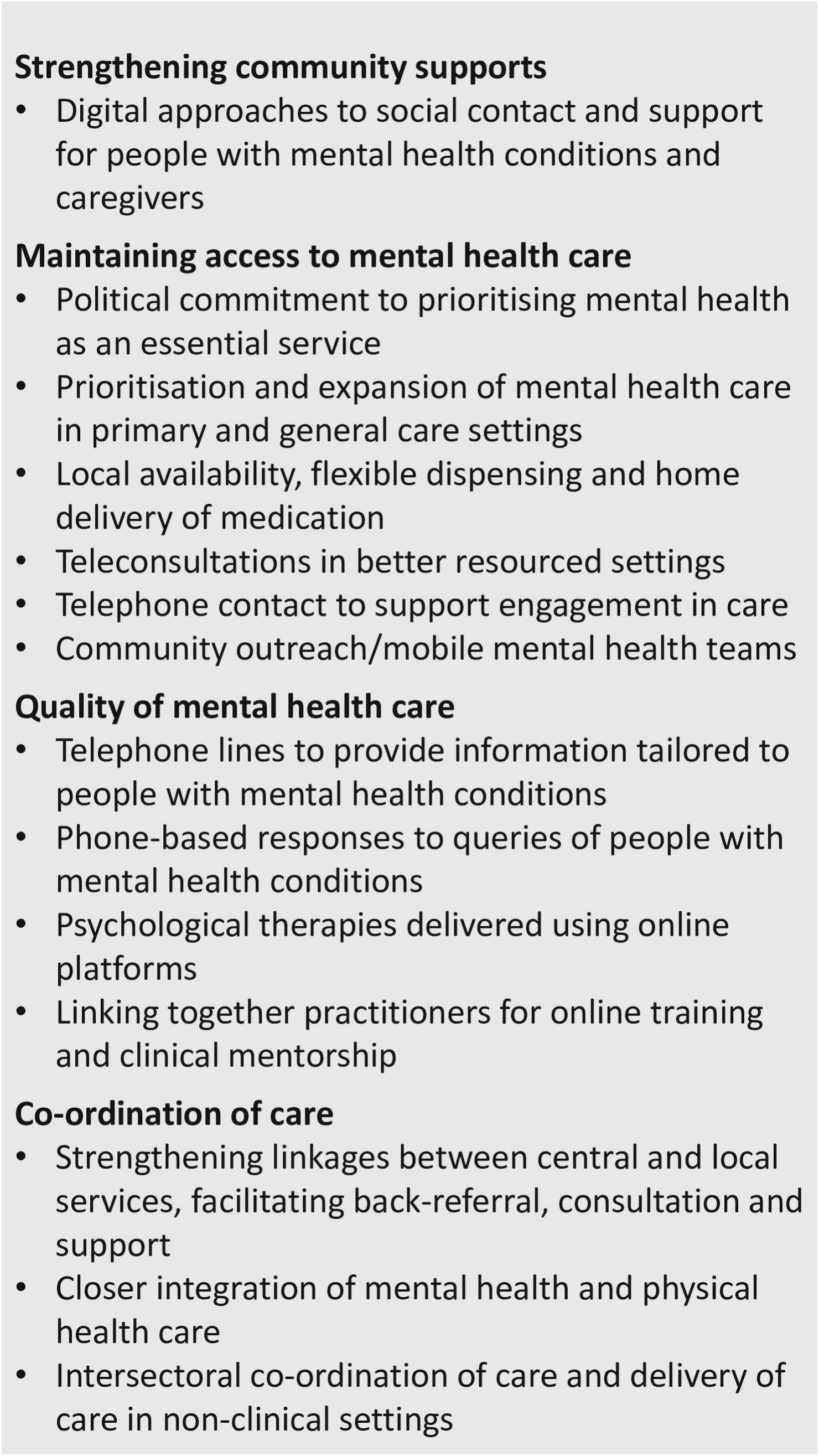
Innovations and strengthening system resilience in response to the pandemic.

## Discussion

In the MASC study, using comparable methods across six countries, we were able to triangulate data from multiple sources and, in several countries, combine with national expert consensus to obtain the most complete picture of impacts of the COVID-19 pandemic on mental health services and service users in LMICs to date. There are several key implications of our findings for increasing preparedness and health system resilience for future disasters.

First, the pernicious effect of mental illness-related stigma was identified in all the study sites and contributed towards the especially low status given by policy makers and health service leaders towards mental health care during the first phase of the pandemic. This is consistent with the findings of the early WHO rapid appraisal of the impact of COVID-19 on mental health care ^6^ and is also fully in line with the main findings of the Lancet Commission on Ending Stigma and Discrimination in Mental Health ^10^. This tendency to provide low rates of funding for mental health care in routine practice, and lower rates still during times of crisis is identified in the Lancet Commission report as ‘structural stigma.’ The consequences of interruptions to mental health services and the stripping away of vital psychosocial support were felt acutely by people with MHCs and their families. Despite WHO calls for mental health care to be designated an essential service to be maintained during the pandemic, this was manifestly not the case on the ground in many countries, particularly for the low-income countries represented in MASC.

Addressing structural stigma requires co-ordinated advocacy from coalitions of stakeholders, but most critically must involve people with lived experience of MHCs and their families and informal caregivers^37^. However, mental health service users are often marginalised, disempowered and may not have strong collective voices where the need is greatest^38^. Research-based efforts have shown that it is possible to equip and empower people with lived experience of MHCs to mobilise, advocate and participate in evidence-based social contact interventions aimed at reducing stigma and increasing commitment of planners and providers to mental health care^39^. However, accountability of governments, resources for nascent service user associations and political investments are needed to make involvement meaningful and sustained.

Second, despite pre-existing vulnerabilities of people with MHCs to poorer physical health and excess mortality compared to the general population^11^, and increased risk of contracting COVID-19, and experiencing greater severity of infection and increased COVID-related mortality^40^, people with MHCs faced more barriers to accessing COVID-19 prevention and treatment programmes than the general population. The physical segregation of mental health care from general health care services, reported in most of the countries participating in MASC, contributed to this injustice, but again stigma and the low priority given to the specific needs of people with MHCs exacerbated exclusion. Our findings accord with reports from diverse global settings that people with severe MHCs were not being given sufficient priority for vaccination against COVID-19^26^.

In MASC, the notable exception was Sri Lanka which had succeeded in ‘building back better’ following the 2005 Tsunami and prioritised the integration of mental health care into primary healthcare services. This integrative approach, advocated by the World Health Organization in the mental health Gap Action Programme (mhGAP)^41^ has many immediate benefits for health systems and populations, including direct strengthening of the health system through horizontal rather than vertical programming, increasing access to mental health care through local availability, reducing exposure to institution-based human rights abuses, improving physical health care of people with severe MHCs^42^ and can contribute to making universal health coverage a reality for one of the most underserved groups^43^.

However, beyond immediate effects, integration also increases health system resilience in the face of humanitarian disasters. Many countries did not have the capacity to adapt once the pandemic hit. Indeed, in Ethiopia, funds were diverted away from efforts to scale-up mental health within primary healthcare settings^32^. This underlines the importance of system preparedness and the need for renewed commitment to decentralised, integrated, community-based mental health services globally^44^.

Third, to varying extents, all sites showed accelerated implementation of digital and remote consulting (See Figure 1), but this also brought the risks of exacerbating inequities – the so-called ‘digital divide’ for access to mental health care^45^. In preparing for future disasters, account needs to be taken of the fact that older people, those with lower levels of education, people with low socio-economic status, rural residents and those with severe MHCs are least likely to be able to benefit from digital ‘solutions’ to accessing mental health care. Providers also need to be equipped with the necessary skills and resources.

Nonetheless, digital technologies may have a role to play in improving disaster response. A conspicuous lack of good quality data on service utilisation was evidenced in all MASC data. Information systems for mental health care are not fit for purpose but present an enormous opportunity to improve system responsivity. With robust safeguards to protect confidentiality, joined up electronic medical record systems and electronic databases of caseloads could be employed to identify people with MHCs who need to be prioritised for pro-active outreach care, home-based delivery of medicines, welfare support and tailored prevention messages. At present, research into digital technologies to improve mental health service planning and improvement in LMICs has been sorely neglected^46^.

Limitations of our study include the lack of comparable quantitative utilisation data across countries, concerns about the accuracy of routine data, and reliance on national level experts who may not have represented the situation fully. However, through our snowballing approach to consult more widely and integration of research and grey literature reports we sought to obtain a comprehensive perspective on COVID-19 impacts.

## Conclusions

All the countries included in this study showed low levels of preparedness for the impacts a pandemic would have on mental health services. Indeed, in most countries, existing systems of mental health care do not allow for adequate mental health care at any time, but especially exposed during a pandemic. Immediate and sustained investment is needed to expand access to mental health care through integration into primary care and community platforms, while also addressing structural stigma and technology gaps that could improve mental health care quality and system resilience.

## Supporting information

COREQ reporting checklist

## Data Availability

Quantitative data on service utilisation are included within supplementary information file 4. Qualitative and narrative data cannot be fully anonymised and made publicly available given the small number of key informants in each country. The data underlying the findings can be obtained from the authors on request.

## Availability of Data and Materials

All data supporting the findings of this study are available within the paper and its Supplementary Information.

## Authors’ contributions

CH and GT conceptualized the design of the study, in collaboration with HL, AA, AAA, RA, OA, AA, TD, WF, OG, LJ, NM, AM, EM, TM, OM, IR, CSA, KS, GSB, OTD, AW, SW and ES. The country leads and their teams led the data collection and analysis AA, AAA, RA, OA, AA, TD, WF, OG, LJ, NM, AM, EM, TM, OM, IR, CSA, KS, GSB, OTD, AW and SW. EB contributed to the statistical and quantitative cross-country analyses and HL led the qualitative cross-country analysis. MM and NV coordinated the study and contributed to the drafting of the manuscript. All authors commented actively along the process, read and approved the final version of the manuscript.

## Acknowledgements

Team activities in Chile received support from the Research Fund of the School of Public Health, University of Chile. In Ethiopia, MASC researchers were supported by the National Institute for Health and Social Care Research (NIHR) Global Health Research Unit on Health System Strengthening in Sub-Saharan Africa, King’s College London (GHRU 16/136/54) using UK aid from the UK Government to support global health research. The views expressed in this publication are those of the authors and not necessarily those of the NIHR or the Department of Health and Social Care or Public Health England. CH, AA, AM and WF were supported by the AMARI project (African Mental Health Research Initiative) as part of the DELTAS (Developing Excellence in Leadership, Training and Science) Africa Initiative [DEL-15-01].

GT is supported by the National Institute for Health and Care Research (NIHR) Applied Research Collaboration South London (NIHR ARC South London) at King’s College Hospital NHS Foundation Trust; CH, GT and AA receive support from NIHR through the NIHR Global Health Research Group on Homelessness and Mental Health in Africa (NIHR134325) and CH also receives support from the SPARK project (NIHR200842), using UK aid from the UK Government. The views expressed are those of the authors and not necessarily those of the NIHR or the Department of Health and Social Care. GT is also supported by the UK Medical Research Council (UKRI) for the Indigo Partnership (MR/R023697/1) awards. CH, AA and WF receive support from the Wellcome Trust through grants 222154/Z20/Z. CH also receives support from WT grant 223615/Z/21/Z. HL is supported by the MRC-UKRI in relation to the Indigo Partnership (MR/R023697/1) and Adolescents’ Resilience and Treatment nEeds for Mental health in Indian Slums (ARTEMIS) Grant no: MR/S023224/1 awards.

OG receives support from the UK Medical Research Council (UKRI) through the International Research Programme on Psychoses in Diverse Settings (INTREPID III) award (MR/X022242/1). We would like to thank the heads of the facilities, mental health services providers and service users that agreed to participate in our study and for those people who supported data collection.

For the purpose of open access, the authors have applied a Creative Commons Attribution (CC BY) licence to any Author Accepted Author Manuscript version arising from this submission.

