## Supplementary material for "Adverse sequelae of the COVID-19 pandemic on mental health care in six low- and middle-income countries: MASC study": COREQ reporting checklist

**Consolidated criteria for reporting qualitative studies (COREQ)**

Developed from: Tong A, Sainsbury P, Craig J. Consolidated criteria for reporting qualitative research (COREQ): a 32-item checklist for interviews and focus groups. *International Journal for Quality in Health Care*. 2007. Volume 19, Number 6: pp. 349 – 357

|  | **Criteria description** | **Study information** | | | | | | **Location in manuscript** |
| --- | --- | --- | --- | --- | --- | --- | --- | --- |
|  |  | **Chile** | **Ethiopia** | **Georgia** | **Nigeria** | **South Africa** | **Sri Lanka** |  |
| **Domain 1: Research team and reflexivity** | | | | | | | | |
| Personal characteristics | | | | | | | | |
| 1. Interviewer/ facilitator | Which author/s conducted the interview or focus group? | OT, CS | AM, WF | TM, LD | OA, AA | TD, KS | AW, SW | Table 1 (page 10) |
| 2. Credentials | What were the researcher’s credentials? E.g. PhD, MD | PhD, MSc | MA, MSc | PhD student;  MA student; | MD | PhD | For AW, SW-  FRCPsych UK  MD Psych Sri Lanka  For AW- MSc | Table 1 (page 10) |
| 3. Occupation | What was their occupation at the time of the study? | Both mental health researchers; mental health professional. | Clinical psychologist; mental health professional | Mental health professionals | Psychiatrist | Registered counsellor, health psychologist | Consultant Psychiatrist/senior lecturer | Table 1 (page 10) |
| 4. Gender | Was the researcher male or female? | Both female | Both male | Both female | Both male | Female | AW- Female,  SW-Male | Table 1 (page 10) |
| 5. Experience and training | What experience or training did the researcher have? | Experienced with qualitative data collection and analysis | Experienced and trained with qualitative data collection and analysis | TM was experienced with qualitative and quantitative data collection and analysis; LD was exposed to qualitative data collection | Experienced with qualitative data collection and analysis | Experienced with qualitative data collection | Experienced with qualitative data collection | Page 13 |
| Relationship with participants | | | | | | | | |
| 6. Relationship established | Was a relationship established prior to study commencement? | Some key informants were known to the researchers | Some key informants were known to the researchers | Researchers did not know the key-informants prior to study; all of them were known to NM as colleagues | Most of the informants were known to the researchers, some were not | All key informants were known to the researchers | For some interviewees | Page 13 |
| 7. Participant knowledge of the interviewer | What did the participants know about the researcher? e.g., personal goals, reasons for doing the research | They knew that they were researchers of the School of Public Health and had access to a document explaining the study's objective, before sending informed consent. | Some were aware that they were PhD students, but working on a different topic. | Key-informants knew about the academic background of researchers | Most of the informants were aware that the researchers were academics and their interest in the work was both professional and academic. | As public mental health researchers | Mainly as psychiatrists who are interested in research | Page 13 |
| 8. Interviewer characteristics | What characteristics were reported about the interviewer/facilitator? e.g., Bias, assumptions, reasons and interests in the research topic | Both are recognized for their previous professional experience in mental health services in Chile and their current academic dedication to researching mental health services in Chile. | Both involved in mental health care provision and efforts to expand access to mental health care in Ethiopia | Researchers had academic interests | Both involved in mental health care provision and efforts to expand access to mental health care in Nigeria | Both involved in public mental health in the coutnry | Both involved in caring for psychiatry patients during COVID pandemic in Sri Lanka | Page 13 |
| **Domain 2: Study design** | | | | | | | | |
| Theoretical framework | | | | | | | | |
| 9. Methodological orientation and theory | What methodological orientation was stated to underpin the study? e.g. grounded theory, discourse analysis, ethnography, phenomenology, content analysis | Template analysis | | | | | | Page 13 |
| Participant selection | | | | | | | | |
| 10. Sampling | How were participants selected? e.g. purposive, convenience, consecutive, snowball | Purposively based on their professional role, with some snowballing to identify people involved in mental health response to COVID-19 | | | | | | Page 13 |
| 11. Method of approach | How were participants approached? e.g. face-to-face, telephone, mail, email | Email | Phone or email | Email was used to invite key-informants | Phone, email and social media (WhatsApp) | Approached via email. | Face to face, virtual platforms | Page 13 |
| 12. Sample size | How many participants were in the study? | 144 key informants | | | | | | Page 10, Table 1 |
| 13. Non-participation | How many people refused to participate or dropped out? Reasons? | One person was not available as they were on leave. | One person dropped out as they travelled abroad and were unable to set up virtual meeting. Another one was not available due to family commitments. | 5 persons; no reasons were given | None | All participants who were approached participated. | Out of the approached potential interviewees, two administrators from the ministry of health did not agree to participate due to their busy schedules | Page 13 |
| Setting | | | | | | | | |
| 14. Setting of data collection | Where was the data collected? e.g., home, clinic, workplace | Virtual | Workplace or virtual | Virtual. ZOOM platform was used to conduct interviews | Virtual | Virtual or workplace | Clinic, wards, department, virtual | Table 1 (page 10) |
| 15. Presence of non-participants | Was anyone else present besides the participants and researchers? | A master's student, as a research assistant. | No | No | No | No | No | Page 13 |
| 16. Description of sample | What are the important characteristics of the sample? e.g., demographic data, date | 18 women; 10 men | 3 women; 15 men | 9 female; 1 male | 6 females and 9 males | 10 Female, 7 Male | Males-18  Females-8 | Table 1 (page 10) |
|  |  | April 2021 | August and September 2021 | December 2020 - January 2021 | June 2021 | June-Sept 2021 | Data collection period |  |
| Data collection | | | | | | | | |
| 17. Interview guide | Were questions, prompts, guides provided by the authors? Was it pilot tested? | Yes, there was a topic guide  Yes, it was pilot tested | | | | | | Page 13 |
| 18. Repeat interviews | Were repeat interviews carried out? If yes, how many? | No | | | | | | Page 13 |
| 19. Audio/visual recording | Did the research use audio or visual recording to collect the data? | Yes – audiorecording | | | | | | Page 13 |
| 20. Field notes | Were ﬁeld notes made during and/or after the interview or focus group? | Yes | | | | | | Page 13 |
| 21. Duration | What was the duration of the interviews or focus group? | 40 minutes to 2 hours | | | | | | Page 10, Table 1 |
| 22. Data saturation | Was data saturation discussed? | No | | | | | | N/A |
| 23. Transcripts returned | Were transcripts returned to participants for comment and/or correction? | No, but there were efforts to present the findings to an expert group before finalising | | | | | | Page 13 |
| **Domain 3: analysis and ﬁndings** | | | | | | | | |
| *Data analysis* | | | | | | | | |
| 24. Number of data coders | How many data coders coded the data? | 2 | 2 | 2 | 2 | 2 | 4 | Page 10, Table 1 |
| 25. Description of the coding tree | Did authors provide a description of the coding tree? | Yes | | | | | | Supplementary file 3 |
| 26. Derivation of themes | Were themes identiﬁed in advance or derived from the data? | Themes were defined ahead of time but modified in response to findings | | | | | | Page 13 |
| 27. Software | What software, if applicable, was used to manage the data? | Not applicable | Opencode | n/a | MAXQDA | Opencode | Opencode | Page 10, Table 1 |
| 28. Participant checking | Did participants provide feedback on the ﬁndings? | Yes, see 23. | | | | | | Page 13 |
| Reporting | | | | | | | | |
| 29. Quotations presented | Were participant quotations presented to illustrate the themes/ﬁndings? Was each quotation identiﬁed? e.g. participant number | Yes quotations are used and the participant identification number is provided | | | | | | Results |
| 30. Data and ﬁndings consistent | Was there consistency between the data presented and the ﬁndings? | Yes – multiple iterations with the country teams to ensure accurate and consistent portrayal of findings | | | | | | Results |
| 31. Clarity of major themes | Were major themes clearly presented in the ﬁndings? | Yes | | | | | | Results |
| 32. Clarity of minor themes | Is there a description of diverse cases or discussion of minor themes? | There is a comparative analysis across the countries | | | | | | Results |
