## Supplemental File 3 for "Adverse sequelae of the COVID-19 pandemic on mental health care in six low- and middle-income countries: MASC study"

**Supplementary File 3: MASC Narrative data synthesis tool**

|  |  | **Data sources** | **Date** |
| --- | --- | --- | --- |
| **1** | **Availability of mental health treatment for people with mental illness** |  |  |
| **1.1** | **What changes are people with mental health conditions experiencing at the community level?** |  |  |
|  | Any evidence of increased stigmatization (evidence of increased stigmatizing attitudes), discrimination (access to aid, food security or poverty interventions) or human rights abuses (e.g. violence towards people with MHCs, restraint of people with MHCs) |  |  |
|  | Impact of physical distancing on social contact, social isolation and abandonment of people with MHCs  Differential access to social media or internet- or phone-based means for social contact. |  |  |
|  | Impact on levels of homelessness in people with mental health conditions.  Impact on homeless people with mental health conditions |  |  |
|  | Differential access to health education messages, information about COVID-19 for people with mental health conditions |  |  |
| **1.2.** | **Changes in access to, or availability of, mental health care delivered within primary health care?** |  |  |
|  | Are mental health care services in primary care being maintained (at all, partially)? Who for (which types of mental health conditions)? Urban and rural primary care facilities?  If available, provide figures on changing numbers of contacts for people with mental health conditions seen in PHC (from routine HMIS). |  |  |
|  | Are essential psychotropic medications still being purchased or is priority being directed to medications/equipment for physical health conditions? |  |  |
| **1.3** | **Changes in access to, and availability of, secondary (specialist) mental health services**  **(primary and general hospital levels in Ethiopia]** |  |  |
|  | Are mental health care services in secondary mental health services being maintained (at all, partially)? Who for (which types of mental health conditions)?  If available, provide figures on changing numbers of contacts for people with mental health conditions seen in PHC (from routine HMIS). |  |  |
|  | Are specialist staff in secondary care settings being redeployed to acute care?  Are rooms that are usually used to deliver out-patient mental health care in secondary care settings being used for COVID-19/other acute care responses? |  |  |
|  | Are essential psychotropic medications still being purchased or is priority being directed to medications/equipment for physical health conditions? |  |  |
| **1.4** | **Changes in access to, and availability of, tertiary (specialist) mental health services)**  **(referral hospitals in regions and national level]** |  |  |
|  | Have any tertiary referral centres been closed? |  |  |
|  | Has there been any reduction in mental health beds available in tertiary settings?/wards converted for use for acute care? |  |  |
|  | Have mental health specialists from tertiary settings been redeployed to acute care (or other care) settings? |  |  |
|  | Availability of essential investigations for monitoring people on long-term psychotropic medications |  |  |
|  | Availability of ECT (when indicated), including availability of adequately qualified mental health staff, anesthetic specialist and oxygen |  |  |

| **1.5** | **Changes in access to, and availability of, psychological treatments** |
| --- | --- |
|  | Are face-to-face psychological treatments being delivered? In what settings? |
|  | Are psychology staff being redeployed/diverted from pre-existing services to the COVID-19 MHPSS response? |
|  | Are psychological therapies being delivered remotely? If so, how does this affect access for those without phone/app access? |
| **2** | **Access to, and availability of, physical health care and prevention/promotion activities for people with mental health conditions** |
| **2.1** | **Community level impact on physical health and help-seeking** |
|  | Access to health promotion/illness prevention information related to COVID-19 for people with mental health conditions (in the community and in mental health facilities) |
|  | Any evidence of less access to hand sanitizer/masks/gloves if prices rise for people with low resources?  Evidence of people with mental health conditions differentially exposed to crowding, shared water and sanitation facilities that may increase risk of COVID-19? |
|  | Any evidence of increased vulnerability of people with mental health conditions to (1) contracting COVID-19 infection and (2) serious/fatal outcomes because of frequency of co-occurring conditions, and services/protection issues outlined above? |
|  | Evidence of lower resources of households of a person with a mental health condition and societal exclusion that may result in them not being able to access treatment and care for COVID-19 or other physical conditions? |
|  | People with mental health conditions less able to access healthcare services due to interruptions in public transport |

| **2.2** | **Access to physical health care in general healthcare settings** |
| --- | --- |
|  | Impact of changing thresholds for severity for services to assess/accept cases of physical illness |
|  | Evidence that people with mental health conditions are less often tested/treated/admitted/treated in the Intensive Care Unit for COVID-19 symptoms  Triage criteria for treatment/ITU that include any criteria related to MI |
|  | Evidence that people with mental health conditions have less access to palliative/end of life care |
| **2.3** | **Access to physical health care in mental health care settings** |
|  | Availability of thermometers |
|  | Levels of testing for COVID-19 in mental health care settings (compared to other hospital settings) |
|  | Access to, and availability of, COVID-19-related treatment in mental health care settings or for people who need to be transferred from mental health care settings (any barriers with hospital transfers?); availability of oxygen |
| **2.4** | **Access to physical health care in social care settings (e.g. institutional NGO care)** |
|  | Availability of thermometers |
|  | Levels of testing for COVID-19 |
|  | Access to, and availability of, COVID-19-related treatment in mental health care settings or for people who need to be transferred from mental health care settings (any barriers with hospital transfers?); availability of oxygen |
