## Supplemental File 2 for "Adverse sequelae of the COVID-19 pandemic on mental health care in six low- and middle-income countries: MASC study"

**Supplementary File 2: MASC rating tool**

| **1** | **Site details, date of assessment and name of person completing form** | |
| --- | --- | --- |
| **2** | **Which of the following interventions/services related to mental, neurological and substance use (MNS) disorders have been disrupted due to COVID-19?** | |
| **2A** | **Management of emergency mental illness presentations**  **(e.g. acute behavioural disturbance, restrained patient)** | |
| 2A(1) | Specialist mental health services | Service operating at usual level |
|  |  | Partially reduced service |
|  |  | Substantially reduced service |
|  |  | No service |
|  |  | Don’t know |
|  |  | No service available pre-COVID-19 |
| 2A(2) | General hospital settings | Service operating at usual level |
|  |  | Partially reduced service |
|  |  | Substantially reduced service |
|  |  | No service |
|  |  | Don’t know |
|  |  | No service available pre-COVID-19 |
| 2A(3) | Primary care level | Service operating at usual level |
|  |  | Partially reduced service |
|  |  | Substantially reduced service |
|  |  | No service |
|  |  | Don’t know |
|  |  | No service available pre-COVID-19 |
| 2A(4) | Private / NGO sectors | Service operating at usual level |
|  |  | Partially reduced service |
|  |  | Substantially reduced service |
|  |  | No service |
|  |  | Don’t know |
|  |  | No service available pre-COVID-19 |

| **2B** | **Management of status epilepticus** | |
| --- | --- | --- |
| 2B(1) | Specialist mental health services | Service operating at usual level |
|  |  | Partially reduced service |
|  |  | Substantially reduced service |
|  |  | No service |
|  |  | Don’t know |
|  |  | No service available pre-COVID-19 |
| 2B(2) | General hospital settings | Service operating at usual level |
|  |  | Partially reduced service |
|  |  | Substantially reduced service |
|  |  | No service |
|  |  | Don’t know |
|  |  | No service available pre-COVID-19 |
| 2B(3) | Primary care level | Service operating at usual level |
|  |  | Partially reduced service |
|  |  | Substantially reduced service |
|  |  | No service |
|  |  | Don’t know |
|  |  | No service available pre-COVID-19 |
| **2C** | **Management of emergency substance use presentations**  **(severe substance withdrawal syndromes)** | |
| 2C(1) | Specialist mental health services | Service operating at usual level |
|  |  | Partially reduced service |
|  |  | Substantially reduced service |
|  |  | No service |
|  |  | Don’t know |
|  |  | No service available pre-COVID-19 |
| 2C(2) | General hospital settings | Service operating at usual level |
|  |  | Partially reduced service |
|  |  | Substantially reduced service |
|  |  | No service |
|  |  | Don’t know |
|  |  | No service available pre-COVID-19 |

| 2C(3) | Primary care | Service operating at usual level |
| --- | --- | --- |
|  |  | Partially reduced service |
|  |  | Substantially reduced service |
|  |  | No service |
|  |  | Don’t know |
|  |  | No service available pre-COVID-19 |
| **2D** | **Ongoing follow-up of people with existing mental health conditions, neurological or substance use disorders** | |
| 2D(1) | Specialist mental health services | Service operating at usual level |
|  |  | Partially reduced service |
|  |  | Substantially reduced service |
|  |  | No service |
|  |  | Don’t know |
|  |  | No service available pre-COVID-19 |
| 2D(2) | General hospital settings | Service operating at usual level |
|  |  | Partially reduced service |
|  |  | Substantially reduced service |
|  |  | No service |
|  |  | Don’t know |
|  |  | No service available pre-COVID-19 |
| 2D(3) | Primary care level | Service operating at usual level |
|  |  | Partially reduced service |
|  |  | Substantially reduced service |
|  |  | No service |
|  |  | Don’t know |
|  |  | No service available pre-COVID-19 |
| **2E** | **Psychotherapy/counselling/psychosocial interventions for mental health conditions or substance use disorders** | |
| 2E(1) | Specialist mental health services | Service operating at usual level |
|  |  | Partially reduced service |
|  |  | Substantially reduced service |
|  |  | No service |
|  |  | Don’t know |
|  |  | No service available pre-COVID-19 |
| 2E(2) | General hospital settings | Service operating at usual level |
|  |  | Partially reduced service |
|  |  | Substantially reduced service |
|  |  | No service |
|  |  | Don’t know |
|  |  | No service available pre-COVID-19 |
| **2E** | **Psychotherapy/counselling/psychosocial interventions for mental health conditions or substance use disorders** | |
| 2E(3) | Primary care level | Service operating at usual level |
|  |  | Partially reduced service |
|  |  | Substantially reduced service |
|  |  | No service |
|  |  | Don’t know |
|  |  | No service available pre-COVID-19 |
| **2F** | **Medications for mental health conditions, neurological or substance use disorders** | |
| 2F(1) | Specialist mental health services | Service operating at usual level |
|  |  | Partially reduced service |
|  |  | Substantially reduced service |
|  |  | No service |
|  |  | Don’t know |
|  |  | No service available pre-COVID-19 |
| 2F(2) | General hospital settings | Service operating at usual level |
|  |  | Partially reduced service |
|  |  | Substantially reduced service |
|  |  | No service |
|  |  | Don’t know |
|  |  | No service available pre-COVID-19 |
| 2F(3) | Primary care level | Service operating at usual level |
|  |  | Partially reduced service |
|  |  | Substantially reduced service |
|  |  | No service |
|  |  | Don’t know |
|  |  | No service available pre-COVID-19 |

| **2G** | **Diagnostic and laboratory services for people with mental health conditions** | |
| --- | --- | --- |
| 2G(1) | Specialist mental health services | Service operating at usual level |
|  |  | Partially reduced service |
|  |  | Substantially reduced service |
|  |  | No service |
|  |  | Don’t know |
|  |  | No service available pre-COVID-19 |
| 2G(2) | General hospital settings | Service operating at usual level |
|  |  | Partially reduced service |
|  |  | Substantially reduced service |
|  |  | No service |
|  |  | Don’t know |
|  |  | No service available pre-COVID-19 |
| 2G(3) | Primary care level | Service operating at usual level |
|  |  | Partially reduced service |
|  |  | Substantially reduced service |
|  |  | No service |
|  |  | Don’t know |
|  |  | No service available pre-COVID-19 |

| **3** | **What are the main reasons for mental health service disruption (for each level of the health system)? Tick all that apply.** | | | |
| --- | --- | --- | --- | --- |
|  |  | Specialist mental health care  (1) | General hospitals  (2) | Primary care  (3) |
| 3A | Closure of services due to regional/zonal/district plans |  |  |  |
| 3B | Decrease in patient volume due to patients not presenting |  |  |  |
| 3C | Insufficient staff to provide services |  |  |  |
| 3D | Mental health trained staff deployed to provide COVID-19 clinical management or emergency support |  |  |  |
| 3E | The clinical set up has been designated as COVID-19 care facility |  |  |  |
| 3F | Insufficient Personal Protective Equipment (PPE) available for health care providers to provide services |  |  |  |
| 3G | Unavailability/stock out of essential medicines |  |  |  |
| 3H | Travel restrictions hindering access to the health facilities for patients |  |  |  |
| 3J | Others (please specify what are the other causes of this disruption): |  |  |  |
| **4** | **What modifications have been made to the delivery of mental health care to overcome disruption/respond to COVID-19 (for each level of the health system)?**  **Tick any that apply.** | | | |
|  |  | Specialist mental health care  (1) | General hospitals  (2) | Primary care  (3) |
| 4A | Lengthening time between OPD visits/ increasing lengths of prescriptions |  |  |  |
| 4B | Arranging for medication to be picked up at local pharmacy outlets/other flexible dispensing arrangements |  |  |  |
| 4C | Telephone or internet-based consultations |  |  |  |
| 4D | Triaging facility-based consultations to focus on emergency presentations |  |  |  |
| 4E | Back-referral to more locally available mental health care |  |  |  |
| 4F | Expanding access to self-help interventions (e.g. through information leaflets, online resources) |  |  |  |
| 4G | Raising community awareness of importance of continuing with mental health care & seeking help for new mental health conditions |  |  |  |
| 4H | Identifying high-risk patients (for relapse/disengagement) for proactive community outreach from community health workers |  |  |  |
| 4J | Identifying patients vulnerable to abandonment/food insecurity and linking with community support |  |  |  |
| 4K | Identifying patients vulnerable to violence/restraint or other human rights abuses and arranging interventions |  |  |  |
| 4L | Integrating COVID-19 health education with mental care services |  |  |  |
| 4M | Other? (specify) |  |  |  |
