## Supplemental File 1 for "Adverse sequelae of the COVID-19 pandemic on mental health care in six low- and middle-income countries: MASC study"

**Supplementary File 1: Topic guide for Semi-Structured Interviews and Focus Group Discussions**

| Country |  |
| --- | --- |
| Date of interview |  |
| Identification number |  |
| Gender |  |
| Type of role | Government (health) |
|  | Government (emergency) |
|  | Government (social/economic) |
|  | Mental health service manager |
|  | Mental health expert by experience |
|  | Non-governmental organisation |
|  | Other (specify) |

1. **What do you think have been the main impacts of the COVID-19 pandemic on people with mental health conditions in [NAME OF COUNTRY]?**

Probes: in terms of their physical health? stigma/discrimination? Human rights abuses? Economic impact? Abandonment? Homelessness? Mental health? Physical health?

Probe for specific examples based on the respondent’s experience.

2. **What has been the impact of COVID-19 on mental health care in [INSERT NAME COUNTRY-SITE]?**

Probes: Service access for mental health care? Service access for physical health care? Redeployment of mental health specialists? Quality of care?

3. **What kinds of modifications have been made to mental health care delivery to overcome barriers to access/quality caused by COVID-19? In secondary and tertiary care? In primary care? In NGO settings?**

Probes: Which of these modifications have been beneficial for improving care delivery? Which modifications would you like to see more widely applied? How might these modifications be useful for improving service delivery post-COVID delivery?

Probe for detailed descriptions of successful service modifications.

4. **How is access to information on COVID-19, testing and treatment for people with mental health conditions compared to those without?**

Probes:

In the community? In mental health service settings? (e.g. in-patient care)

What mental health support is provided in COVID-19 treatment and isolation centres? How do COVID-19 treatment centres manage people with mental health conditions? What are the challenges? What has been done well?

Probe for specific examples of inequities experienced by people with mental health conditions.

5. **How are the infection prevention and control measures for COVID-19 being implemented in mental health care settings? How does this compare to other medical settings?**

Probe for examples of inequities.

6. **What interventions have been made to mitigate the impact of COVID-19 infection prevention and control measures (e.g. social distancing, isolation) on people with mental health conditions?**

Probes: How adequate have these measures been? To what extent do you think people with mental health conditions are excluded from these mitigation interventions? (e.g. food supplements, financial support, social support). In your view, what else could be done?

Probe for detailed descriptions of effective interventions.

7. **How do you think the rights of people with mental health conditions have been affected by COVID-19?**

Probe: in terms of policy changes? Legislation changes? Service delivery changes? Changes in community attitudes?
